# Performance assessment of 11 commercial serological tests for SARS-CoV-2 on hospitalized COVID-19 patients

**DOI:** 10.1101/2020.08.06.20168856

**Authors:** C Serre-Miranda, C Nobrega, S Roque, J Canto-Gomes, CS Silva, N Vieira, P Barreira-Silva, P Alves-Peixoto, J Cotter, A Reis, M Formigo, H Sarmento, O Pires, A Carvalho, DY Petrovykh, L Diéguez, JC Sousa, N Sousa, C Capela, JA Palha, PG Cunha, M Correia-Neves

## Abstract

Commercial availability of serological tests to evaluate immunoglobulins (Ig) towards severe acute respiratory syndrome coronavirus 2 (SARS-CoV-2) has grown exponentially since the onset of COVID-19 (Coronavirus Disease 2019) outbreak. Their thorough validation is of extreme importance before using them as epidemiological tools to infer population seroprevalence, and as complementary diagnostic tools to molecular approaches *(e.g*. RT-qPCR). Here we assayed commercial serological tests (semiquantitative and qualitative) from 11 suppliers in 126 samples collected from hospitalized COVID-19 patients, and from 36 healthy and HIV-infected individuals (collected at the pre-COVID-19 pandemic). Specificity was above 95% in 9 tests. Samples from COVID-19 patients were stratified by days since symptoms onset (<10, 10-15, 16-21 and >21 days). Tests sensitivity increases with time since symptoms onset, and peaks at 16-21 days for IgM and IgA (maximum: 91.2%); and from 16-21 to >21 days for IgG, depending on the test (maximum: 94.1%). Data from semiquantitative tests show that patients with severe clinical presentation have lower relative levels of IgM, IgA and IgG at <10 days since symptoms onset in comparison to patients with non-severe presentation. At >21 days since symptoms onset the relative levels of IgM and IgG (in one test) are significantly higher in patients with severe clinical presentation, suggesting a delay in the upsurge of Ig against SARS-CoV-2 in those patients.

This study highlights the high specificity of most of the evaluated tests, and sensitivity heterogeneity. Considering the virus genetic evolution and population immune response to it, continuous monitoring of commercially available serological tests towards SARS-CoV-2 is necessary.

## INTRODUCTION

The severe acute respiratory syndrome coronavirus 2 (SARS-CoV-2) is a large RNA virus from the *Coronaviridae* virus family that is currently globally spread (1,2). Considering the absence of an effective vaccine or treatment for SARS-CoV-2 infection, early diagnosis of infection and isolation of infected individuals is critical to control the ongoing pandemic (3). Most efforts for case detection involve collection of swab samples from the upper respiratory tract, and the amplification of viral nucleic acids sequences by RT-qPCR. These sequences include genes encoding for the viral proteins: envelop (E), RdRp, nucleocapsid (N) 1 and 2, and spike (S) (4). However, RT-qPCR-based diagnosis is time-consuming, expensive and requires highly trained professionals. Serological tests arise as interesting complementary diagnostic tools, but also as means to detect the presence of antibodies towards SARS-CoV-2 at the population level.

Following SARS-CoV-2 infection, most patients produce detectable immunoglobulins (Ig) against a set of viral antigens, particularly to the immunodominant N and S proteins (5-8). Current evidence suggests that Ig produced against these antigens may confer protection against SARS-CoV-2 infection (9-11). Nevertheless, there is still insufficient data on the timing of Ig production upon infection. Literature suggests that, considering the timing since symptoms onset, blood IgA and IgM are detected after 6-8 days; IgA increases continuously up to 20-22 days, and IgM peaks at 10-14 days (5,12-15). IgG seroconversion seems to occur slightly later than IgM, at 9-10 days since symptoms onset. However, many patients seroconvert for both IgM and IgG simultaneously, peaking at around 21 days (13,16-18). Duration and magnitude of Ig response likely correlates with disease severity, and it is yet debatable whether Ig levels remain at sufficient protective levels for long periods of time after viral clearance (17,18).

Serological studies have the potential to help in understand individual and herd immunity to a viral infection (19). Several studies evaluated and compared the performance of serological assays (12,20,21). Notably, the performance (sensitivity and specificity) of these assays can be affected by many variables including: timing of assessment since symptoms/infection, course of COVID-19 (Coronavirus Disease 2019) (from asymptomatic to lethal) and, potentially, population and virus genetics (22,23). It is thus unequivocally important to evaluate the performance of available serological tests in distinct populations and countries, to be able to select the most adequate tests.

Herein, we evaluate the performance of serological tests (3 semiquantitative and 8 qualitative) from 11 suppliers using plasma samples of hospitalized patients with COVID-19 from the Minho region, in the North of Portugal. These tests were chosen considering previous reports on their sensitivity, specificity, and availability.

## METHODS

### Study population

Patients living in the Minho region of Portugal, followed-up as inpatients at Senhora da Oliveira Hospital (Guimarães) and Braga Hospital, diagnosed with COVID-19 (by RT-qPCR at a reference laboratory; at least two positive RT-qPCR results were obtained from each patient) were invited to participate in the study. This study was approved by the Ethics Committees of both participating Hospitals (Senhora da Oliveira Hospital: 25/2020; Braga Hospital: 37/2020). An explanation of the project was provided to those individuals, and the ones that agreed to participate signed an informed consent form. The informed consent was prepared according to the Declaration of Helsinki principles, the Oviedo Convention and according to the General Data Protection Regulation – Regulation (EU) 2016/679. Patients’ blood samples were collected throughout their hospitalization, at different timepoints following symptoms onset. The number of samples available from each participant vary depending on the duration of their hospitalization.

For sensitivity calculation, COVID-19 patients were stratified based on the number of days since symptoms onset as follows: <10 days; 10 to 15 days; 16 to 21 days; >21 days. Days since symptoms onset were calculated based on each patient’s self-report of symptoms manifestation. COVID-19 patients were further categorized according to the severity of their clinical presentation. Patients given oxygen therapy above 10 L/min and/or needing mechanical ventilation (non-invasive or invasive) were considered as having a severe clinical presentation. All other patients (needing supplementary oxygen therapy below 10 L/min and not requiring mechanical ventilator support) were considered as having non-severe clinical presentation. SARS-CoV-2 non-infected controls were selected from banked human plasma samples from two studies at pre-COVID-19 pandemic (the first COVID-19 case in Portugal was reported in early March 2020): i) a study with healthy individuals older than 55 years of age (samples collected between April 2019 and January 2020); ii) a study with HIV-infected patients on antiretroviral therapy for 54 to 60 months (samples collected between January 2016 and August 2018) (24). In both cases, matched samples were selected based on individuals’ sex and age. Control samples were collected, processed and preserved at -80 °C using a similar protocol as the one used for samples from COVID-19 inpatients (bellow).

Data was handled anonymously. Individuals’ sex, age and comorbidities are summarized in Table 1.

**Table 1.** Clinical and demographic characterization of the cohort.

|  | COVID-19 patients |  |  | Pre-COVID-19 controls |  |
| --- | --- | --- | --- | --- | --- |
|  | All | Clinical presentation |  | Healthy controls | HIV and other viral infections |
|  |  | Severe<sup>1</sup> | Non-severe<sup>1</sup> |  |  |
| ***n*** | 89 | 32 | 57 | 25 | 11 |
| **Age (years)**<br>Median [min;max] | 71 [30;96] | 75 [45;96] | 67 [30;94] | 71 [59;80] | 57 [33;72] |
| **Female, *n* (%)** | 51 (57.3) | 16 (50.0) | 35 (61.4) | 13 (52.0) | 3 (27.3) |
| **Hypertension, *n* (%)** | 59 (66.3) | 25 (78.1) | 34 (59.6) | n/a | n/a |
| **Diabetes, *n* (%)** | 27 (30.3) | 11 (34.4) | 16 (28.1) | n/a | n/a |
| **Neoplasia, *n* (%)** | 8 (9.0) | 4 (12.5) | 4 (7.0) | n/a | n/a |
| **Autoimmune disease, *n* (%)** | 5 (5.6) | 2 (6.3) | 3 (5.3) | n/a | n/a |
| **Immunosuppressive drugs, *n* (%)** | 5 (5.6) | 3 (9.4) | 2 (3.5) | n/a | n/a |

<sup>1</sup> Please see "Study Population" section on Methods. n/a: not available

### Sample processing

From each patient, venous blood was collected into K_2_EDTA collecting tubes and processed on the same day: blood collecting tubes were centrifuged at 2000g for 15 min, at 20 °C. Plasma was aliquoted into screw-cap tubes and frozen at -80°C until tested.

### Immunoassays

Semiquantitative [enzyme linked immune-absorbent assays (ELISA) and chemiluminescence immunoassays (CLIA)] and qualitative assays [lateral flow immunoassays (LFIA)] from 11 different suppliers were tested according to manufacturer’s instructions (Supplementary Table S1). At least two different tests were performed for each sample (Supplementary Table S2, S3 and S4).

### Data analysis

For each test, specificity was calculated as the percentage of negative tests among the pre-COVID-19 controls, and sensitivity as the percentage of positive tests among the SARS-CoV-2 confirmed cases. Sensitivity was calculated upon stratification in days since symptoms onset (<10, 10-15; 16-21 and >21 days). Positive predictive value (PPV) was calculated as the proportion of true positive cases *(i.e*. positive serological test on confirmed SARS-CoV-2 infection by RT-qPCR) among the total positive tests, and the negative predictive value (NPV) as the proportion of true negative cases *(i.e*. negative serological test on pre-COVID-19 pandemic samples) among the total negative tests. Whenever the same patient was tested multiple times within the same time range during the course of the disease, only one plasma sample was considered to calculate sensitivity, PPV and NPV (Supplementary Table S2). The 95% confidence intervals for sensitivity, specificity, PPV and NPV were calculated using the Wilson score with continuity correction method. Tests agreement was evaluated using Cohen’s Kappa. For the semiquantitative tests, receiver-operator characteristic (ROC) curves were constructed and used to calculate the area under the curve (AUC) of the different serologic tests. All variables analysed had a non-normal distribution, as verified by Shapiro-Wilk tests. Comparisons of the relative amounts of Ig at the various time ranges were performed using the Kruskal-Wallis test, followed by Dunn’s post-hoc tests. Comparisons of the relative amount of Ig in the groups of patients with severe and non-severe clinical presentation were performed using the Mann-Whitney U test. Differences were considered significant when *p* <0.05. Statistical analyses were performed on GraphPad Prism version 8.4.3 (La Jolla, San Diego, CA, USA).

## RESULTS

### Study population

This study includes 89 hospitalized infected with SARS-CoV-2 (diagnosed by RT-qPCR). Plasma samples from 49 individuals were tested at a single timepoint, whereas 40 individuals were tested more than once during hospitalization (Supplementary Table S2 and S4). Most participants are women (57%; Table 1) with a median age of 71. None of the patients is positive for HIV nor was submitted to previous organ transplantation.

### Performance of tests

Serologic tests from 11 suppliers were assayed for their performance. Specificity considering IgM and IgG combined range from 76.0% (Cellex) to 100.0% (Liming and Render). Considering each Ig class independently, 4 out of 10 tests assessing IgG, and 5 out of 9 tests assessing IgA or IgM show 100.0% specificity (Figure 1 and Table 2). Thirty-six samples were used as negative controls (collected during pre-COVID-19 pandemic); 10 negative controls are positive for at least 1 test; 5 of which tested positive in more than 1 test (Supplementary Table S3). As for the semiquantitative tests (Abbott, Euroimmun and Snibe), 2 of the negative control samples test positive in the Snibe IgG test and have a value close to the lower detection limit (1.124 and 1.652 AU/mL; cut-off 1.000 AU/mL; Supplementary Figure S1).

**Figure 1.**
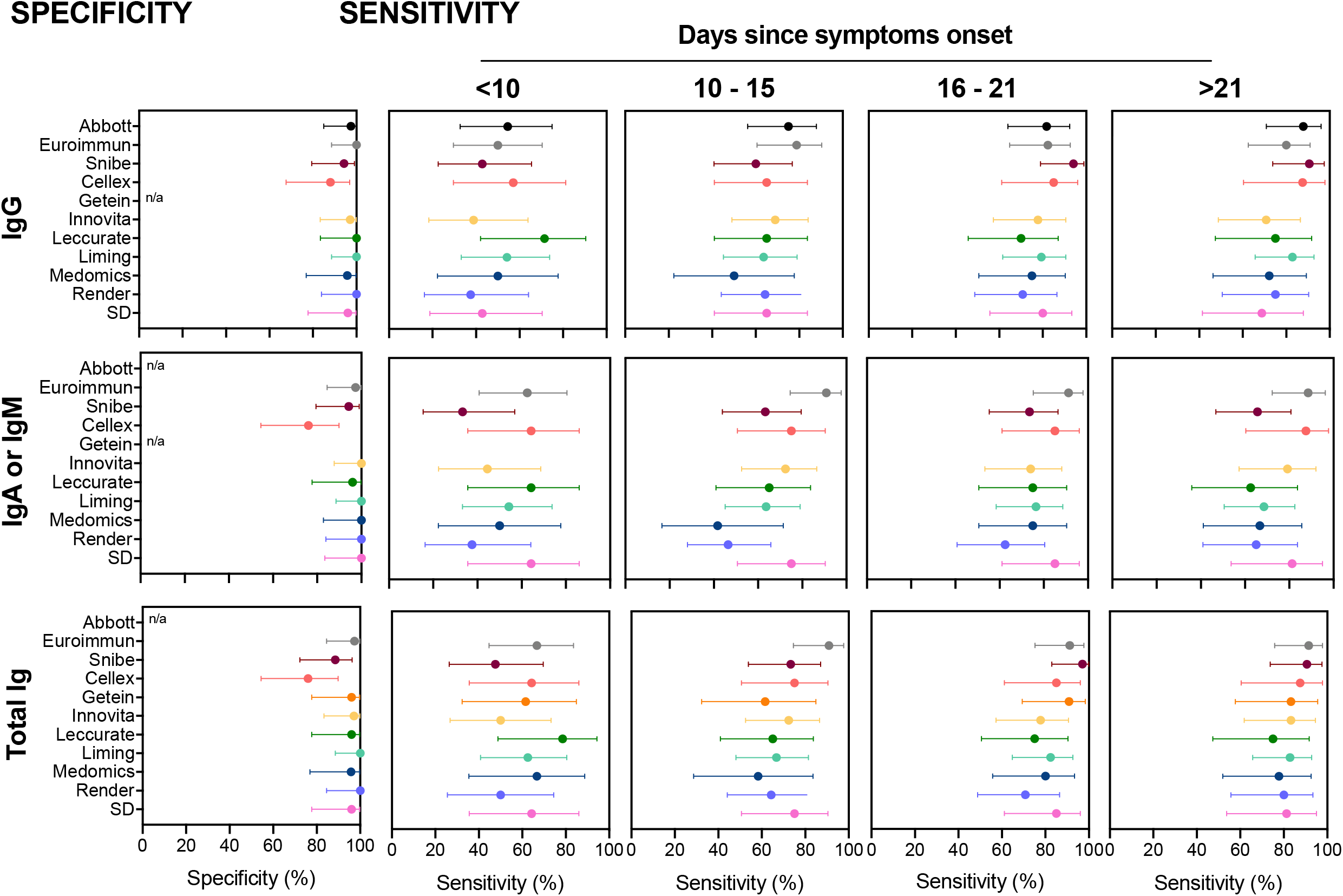
Specificity and sensitivity of the assayed tests to detect immunoglobulins specific for SARS-CoV-2 infection. Sensitivity was evaluated in COVID-19 patients upon stratification by days since symptoms onset. Each dot represents the specificity or sensitivity, and bars represent the 95% confidence interval. n/a: not available. Ig: immunoglobulin.

**Table 2.** Sensitivity and specificity of the assayed tests to detect immunoglobulins specific for SARS-CoV-2 infection. Sensitivity was evaluated in COVID-19 patients upon stratification by days since symptoms onset.

|  |  | SPECIFICITY |  |  |  |  | SENSITIVITY |  |  |  |  |  |  |  |  |  |  |  |  |  |  |  |  |  |  |  |
| --- | --- | --- | --- | --- | --- | --- | --- | --- | --- | --- | --- | --- | --- | --- | --- | --- | --- | --- | --- | --- | --- | --- | --- | --- | --- | --- |
|  |  |  |  |  |  |  | Days since symptoms onset |  |  |  |  |  |  |  |  |  |  |  |  |  |  |  |  |  |  |  |
|  |  | Tot<br>*n* | Neg<br>*n* | Spec<br>% | 95% CI |  | <10 |  |  |  |  | 10 - 15 |  |  |  |  | 16 - 21 |  |  |  |  | >21 |  |  |  |  |
| Commercial kit |  |  |  |  | Lower | Upper | Tot<br>*n* | Pos<br>*n* | Sens<br>% | 95% CI |  | Tot<br>*n* | Pos<br>*n* | Sens<br>% | 95% CI |  | Tot<br>*n* | Pos<br>*n* | Sens<br>% | 95% CI |  | Tot<br>*n* | Pos<br>*n* | Sens<br>% | 95% CI |  |
| Abbott | IgG | 39 | 38 | 97.4 | 84.9 | 99.9 | 22 | 12 | 54.5 | 32.7 | 74.9 | 32 | 24 | 75.0 | 56.2 | 87.9 | 33 | 27 | 81.8 | 63.9 | 92.4 | 33 | 29 | 87.9 | 70.9 | 96.0 |
| Euroimmun | IgG | 38 | 38 | 100.0 | 88.6 | 100.0 | 24 | 12 | 50.0 | 29.6 | 70.4 | 33 | 26 | 78.8 | 60.6 | 90.4 | 34 | 28 | 82.4 | 64.8 | 92.6 | 35 | 28 | 80.0 | 62.5 | 90.9 |
| Snibe | IgG | 35 | 33 | 94.3 | 79.5 | 99.0 | 21 | 9 | 42.9 | 22.6 | 65.6 | 30 | 18 | 60.0 | 40.7 | 76.8 | 34 | 32 | 94.1 | 78.9 | 99.0 | 32 | 29 | 90.6 | 73.8 | 97.5 |
| Cellex | IgG | 25 | 22 | 88.0 | 67.7 | 96.8 | 14 | 8 | 57.1 | 29.6 | 81.2 | 20 | 13 | 65.0 | 40.9 | 83.7 | 20 | 17 | 85.0 | 61.1 | 96.0 | 16 | 14 | 87.5 | 60.4 | 97.8 |
| Getein | --- | --- | --- | --- | --- | --- | --- | --- | --- | --- | --- | --- | --- | --- | --- | --- | --- | --- | --- | --- | --- | --- | --- | --- | --- |  |
| Innovita | IgG | 35 | 34 | 97.1 | 83.4 | 99.9 | 18 | 7 | 38.9 | 18.3 | 63.9 | 29 | 20 | 69.0 | 49.0 | 84.0 | 27 | 21 | 77.8 | 57.3 | 90.6 | 24 | 17 | 70.8 | 48.8 | 86.6 |
| Leccurate | IgG | 25 | 25 | 100.0 | 83.4 | 100.0 | 14 | 10 | 71.4 | 42.0 | 90.4 | 20 | 13 | 65.0 | 40.9 | 83.7 | 20 | 14 | 70.0 | 45.7 | 87.2 | 16 | 12 | 75.0 | 47.4 | 91.7 |
| Liming | IgG | 38 | 38 | 100.0 | 88.6 | 100.0 | 24 | 13 | 54.2 | 33.2 | 73.8 | 33 | 21 | 63.6 | 45.1 | 79.0 | 34 | 27 | 79.4 | 61.6 | 90.7 | 35 | 29 | 82.9 | 65.7 | 92.8 |
| Medomics | IgG | 24 | 23 | 95.8 | 76.9 | 99.8 | 12 | 6 | 50.0 | 22.3 | 77.7 | 12 | 6 | 50.0 | 22.3 | 77.7 | 20 | 15 | 75.0 | 50.6 | 90.4 | 18 | 13 | 72.2 | 46.4 | 89.3 |
| Render | IgG | 26 | 26 | 100.0 | 84.0 | 100.0 | 16 | 6 | 37.5 | 16.3 | 64.1 | 28 | 18 | 64.3 | 44.1 | 80.7 | 24 | 17 | 70.8 | 48.8 | 86.6 | 20 | 15 | 75.0 | 50.6 | 90.4 |
| SD | IgG | 25 | 24 | 96.0 | 77.7 | 99.8 | 14 | 6 | 42.9 | 18.8 | 70.4 | 20 | 13 | 65.0 | 40.9 | 83.7 | 20 | 16 | 80.0 | 55.7 | 93.4 | 16 | 11 | 68.8 | 41.5 | 87.9 |
| Abbott | --- | --- | --- | --- | --- | --- | --- | --- | --- | --- | --- | --- | --- | --- | --- | --- | --- | --- | --- | --- | --- | --- | --- | --- | --- |  |
| Euroimmun | IgA | 38 | 37 | 97.4 | 84.6 | 99.9 | 24 | 15 | 62.5 | 40.8 | 80.5 | 33 | 30 | 90.9 | 74.5 | 97.6 | 34 | 31 | 91.2 | 75.2 | 97.7 | 35 | 31 | 88.6 | 72.3 | 96.3 |
| Snibe | IgM | 35 | 33 | 94.3 | 79.5 | 99.0 | 21 | 7 | 33.3 | 15.5 | 56.9 | 30 | 19 | 63.3 | 43.9 | 79.5 | 34 | 25 | 73.5 | 55.3 | 86.5 | 32 | 21 | 65.6 | 46.8 | 80.8 |
| Cellex | IgM | 25 | 19 | 76.0 | 54.5 | 89.8 | 14 | 9 | 64.3 | 35.6 | 86.0 | 20 | 15 | 75.0 | 50.6 | 90.4 | 20 | 17 | 85.0 | 61.1 | 96.0 | 16 | 14 | 87.5 | 60.4 | 97.8 |
| Getein | --- | --- | --- | --- | --- | --- | --- | --- | --- | --- | --- | --- | --- | --- | --- | --- | --- | --- | --- | --- | --- | --- | --- | --- | --- |  |
| Innovita | IgM | 35 | 35 | 100.0 | 87.7 | 100.0 | 18 | 8 | 44.4 | 22.4 | 68.7 | 29 | 21 | 72.4 | 52.5 | 86.6 | 27 | 20 | 74.1 | 53.4 | 88.1 | 24 | 19 | 79.2 | 57.3 | 92.1 |
| Leccurate | IgM | 25 | 24 | 96.0 | 77.7 | 99.8 | 14 | 9 | 64.3 | 35.6 | 86.0 | 20 | 13 | 65.0 | 40.9 | 83.7 | 20 | 15 | 75.0 | 50.6 | 90.4 | 16 | 10 | 62.5 | 35.9 | 83.7 |
| Liming | IgM | 38 | 38 | 100.0 | 88.6 | 100.0 | 24 | 13 | 54.2 | 33.2 | 73.8 | 33 | 21 | 63.6 | 45.1 | 79.0 | 34 | 26 | 76.5 | 58.4 | 88.6 | 35 | 24 | 68.6 | 50.6 | 82.6 |
| Medomics | IgM | 24 | 24 | 100.0 | 82.8 | 100.0 | 12 | 6 | 50.0 | 22.3 | 77.7 | 12 | 5 | 41.7 | 16.5 | 71.4 | 20 | 15 | 75.0 | 50.6 | 90.4 | 18 | 12 | 66.7 | 41.2 | 85.6 |
| Render | IgM | 26 | 26 | 100.0 | 84.0 | 100.0 | 16 | 6 | 37.5 | 16.3 | 64.1 | 28 | 13 | 46.4 | 28.0 | 65.8 | 24 | 15 | 62.5 | 40.8 | 80.5 | 20 | 13 | 65.0 | 40.9 | 83.7 |
| SD | IgM | 25 | 25 | 100.0 | 83.4 | 100.0 | 14 | 9 | 64.3 | 35.6 | 86.0 | 20 | 15 | 75.0 | 50.6 | 90.4 | 20 | 17 | 85.0 | 61.1 | 96.0 | 16 | 13 | 81.3 | 53.7 | 95.0 |
| Abbott | --- | --- | --- | --- | --- | --- | --- | --- | --- | --- | --- | --- | --- | --- | --- | --- | --- | --- | --- | --- | --- | --- | --- | --- | --- |  |
| Euroimmun | (IgG;IgA) | 38 | 37 | 97.4 | 84.6 | 99.9 | 24 | 16 | 66.7 | 44.7 | 83.6 | 33 | 30 | 90.9 | 74.5 | 97.6 | 34 | 31 | 91.2 | 75.2 | 97.7 | 35 | 32 | 91.4 | 75.8 | 97.8 |
| Snibe | (IgG;IgM) | 35 | 31 | 88.6 | 72.3 | 96.3 | 21 | 10 | 47.6 | 26.4 | 69.7 | 30 | 22 | 73.3 | 53.8 | 87.0 | 34 | 33 | 97.1 | 82.9 | 99.8 | 32 | 29 | 90.6 | 73.8 | 97.5 |
| Cellex | (IgG;IgM) | 25 | 19 | 76.0 | 54.5 | 89.8 | 14 | 9 | 64.3 | 35.6 | 86.0 | 20 | 15 | 75.0 | 50.6 | 90.4 | 20 | 17 | 85.0 | 61.1 | 96.0 | 16 | 14 | 87.5 | 60.4 | 97.8 |
| Getein | (Total Ig) | 25 | 24 | 96.0 | 77.7 | 99.8 | 13 | 8 | 61.5 | 32.3 | 84.9 | 13 | 8 | 61.5 | 32.3 | 84.9 | 22 | 20 | 90.9 | 69.4 | 98.4 | 18 | 15 | 83.3 | 57.7 | 95.6 |
| Innovita | (IgG;IgM) | 35 | 34 | 97.1 | 83.4 | 99.9 | 18 | 9 | 50.0 | 26.8 | 73.2 | 29 | 21 | 72.4 | 52.5 | 86.6 | 27 | 21 | 77.8 | 57.3 | 90.6 | 24 | 20 | 83.3 | 61.8 | 94.5 |
| Leccurate | (IgG;IgM) | 25 | 24 | 96.0 | 77.7 | 99.8 | 14 | 11 | 78.6 | 48.8 | 94.3 | 20 | 13 | 65.0 | 40.9 | 83.7 | 20 | 15 | 75.0 | 50.6 | 90.4 | 16 | 12 | 75.0 | 47.4 | 91.7 |
| Liming | (IgG;IgM) | 38 | 38 | 100.0 | 88.6 | 100.0 | 24 | 15 | 62.5 | 40.8 | 80.5 | 33 | 22 | 66.7 | 48.1 | 81.4 | 34 | 28 | 82.4 | 64.8 | 92.6 | 35 | 29 | 82.9 | 65.7 | 92.8 |
| Medomics | (IgG;IgM) | 24 | 23 | 95.8 | 76.9 | 99.8 | 12 | 8 | 66.7 | 35.4 | 88.7 | 12 | 7 | 58.3 | 28.6 | 83.5 | 20 | 16 | 80.0 | 55.7 | 93.4 | 18 | 14 | 77.8 | 51.9 | 92.6 |
| Render | (IgG;IgM) | 27 | 27 | 100.0 | 84.5 | 100.0 | 16 | 8 | 50.0 | 25.5 | 74.5 | 28 | 18 | 64.3 | 44.1 | 80.7 | 24 | 17 | 70.8 | 48.8 | 86.6 | 20 | 16 | 80.0 | 55.7 | 93.4 |
| SD | (IgG;IgM) | 25 | 24 | 96.0 | 77.7 | 99.8 | 14 | 9 | 64.3 | 35.6 | 86.0 | 20 | 15 | 75.0 | 50.6 | 90.4 | 20 | 17 | 85.0 | 61.1 | 96.0 | 16 | 13 | 81.3 | 53.7 | 95.0 |

CI: confidence interval; Ig: immunoglobulin; Neg: negative; Pos: positive; Sens: sensitivity; Spec: specificity; Tot: total.

To analyse sensitivity, samples were stratified according to time since symptoms onset (<10, 10-15, 16-21 and >21 days). For each Ig, the lowest sensitivities are observed at <10 days since symptoms onset: the Leccurate IgG test presents the highest sensitivity (71.4%) and Snibe IgM the lowest (33.3%; Figure 1 and Table 2). Detection of IgA or IgM reached a maximum sensitivity at 16-21 days in 7 of the 9 tests performed. The maximum sensitivity for IgG peaked at 16-21 days in half of the tests; the other half at >21 days (Abbott, Cellex, Innovita, Leccurate, and Render). As for Getein, that detects total Ig towards SARS-CoV-2, the peak detection is at 16-21 days. Considering the combined Ig classes, at >21 days, the Euroimmun test (IgA and IgG) provides the highest sensitivity (91.4%), followed by the Snibe test (IgM and IgG; 90.6%). The combined IgM and IgG Leccurate test provides the lowest sensitivity value (75.0%).

Interestingly, the two tests with the overall highest sensitivity (Euroimmun and Snibe) show a moderate agreement index (Cohens’ Kappa) of 0.723 (Table 3). Euroimmun has the best agreement with SD (Cohens’ Kappa = 0.827), and Snibe correlates the best with Getein (Cohens’ Kappa = 0.874). For semiquantitative tests, most of the samples from COVID-19 patients with contradicting results present values far from the cut-off values (Supplementary Figure S1).

**Table 3.** Cohen’s kappa coefficient for the strength of agreement between assayed tests to detect immunoglobulins specific for SARS-CoV-2 infection.

|  |  |  | SD |  | Render |  | Medomics |  | Liming |  | Leccurate |  | Innovita |  | Getein |  | Cellex |  | Snibe |  | Euroimmun |  |
| --- | --- | --- | --- | --- | --- | --- | --- | --- | --- | --- | --- | --- | --- | --- | --- | --- | --- | --- | --- | --- | --- | --- |
|  |  |  | Test result (n) |  | Test result (n) |  | Test result (n) |  | Test result (n) |  | Test result (n) |  | Test result (n) |  | Test result (n) |  | Test result (n) |  | Test result (n) |  | Test result (n) |  |
|  |  |  | - | + | - | + | - | + | - | + | - | + | - | + | - | + | - | + | - | + | - | + |
| Abbott | Test | - | 40 | 6 | 52 | 3 | 39 | 5 | 29 | 7 | 39 | 5 | 61 | 3 | 40 | 7 | 35 | 11 | 57 | 11 | 56 | 17 |
|  | result (n) | + | 0 | 51 | 8 | 57 | 4 | 42 | 9 | 89 | 4 | 42 | 4 | 70 | 3 | 46 | 0 | 51 | 3 | 88 | 5 | 94 |
|  | Cohen's Kappa |  | 0.875 |  | 0.817 |  | 0.800 |  | 0.701 |  | 0.800 |  | 0.898 |  | 0.791 |  | 0.770 |  | 0.817 |  | 0.732 |  |
|  | 95% Lower |  | 0.789 |  | 0.733 |  | 0.700 |  | 0.615 |  | 0.700 |  | 0.832 |  | 0.694 |  | 0.671 |  | 0.747 |  | 0.659 |  |
|  | CI Upper |  | 0.931 |  | 0.879 |  | 0.874 |  | 0.776 |  | 0.874 |  | 0.941 |  | 0.865 |  | 0.847 |  | 0.872 |  | 0.796 |  |
| Euroimmun | Test | - | 34 | 2 | 44 | 0 | 34 | 3 | 22 | 4 | 34 | 3 | 52 | 1 | 36 | 2 | 28 | 8 | 50 | 9 |  |  |
|  | result (n) | + | 6 | 55 | 17 | 60 | 10 | 44 | 20 | 105 | 10 | 44 | 14 | 72 | 8 | 51 | 7 | 54 | 13 | 103 |  |  |
|  | Cohen's Kappa |  | 0.827 |  | 0.720 |  | 0.713 |  | 0.552 |  | 0.713 |  | 0.782 |  | 0.790 |  | 0.667 |  | 0.723 |  |  |  |
|  | 95% Lower |  | 0.734 |  | 0.629 |  | 0.607 |  | 0.469 |  | 0.607 |  | 0.702 |  | 0.693 |  | 0.563 |  | 0.650 |  |  |  |
|  | CI Upper |  | 0.894 |  | 0.796 |  | 0.800 |  | 0.632 |  | 0.800 |  | 0.845 |  | 0.863 |  | 0.757 |  | 0.787 |  |  |  |
| Snibe | Test result (n) | - | 33 | 4 | 44 | 1 | 35 | 3 | 28 | 4 | 35 | 3 | 54 | 1 | 38 | 0 | 31 | 6 |  |  |  |  |
|  |  | + | 7 | 52 | 16 | 58 | 9 | 44 | 11 | 97 | 9 | 44 | 12 | 71 | 6 | 53 | 4 | 55 |  |  |  |  |
|  | Cohen's Kappa |  | 0.762 |  | 0.715 |  | 0.735 |  | 0.718 |  | 0.735 |  | 0.810 |  | 0.874 |  | 0.778 |  |  |  |  |  |
|  | 95% Lower |  | 0.662 |  | 0.624 |  | 0.630 |  | 0.635 |  | 0.630 |  | 0.732 |  | 0.787 |  | 0.679 |  |  |  |  |  |
|  | CI Upper |  | 0.840 |  | 0.792 |  | 0.819 |  | 0.789 |  | 0.819 |  | 0.870 |  | 0.930 |  | 0.854 |  |  |  |  |  |
| Cellex | Test | - | 34 | 1 | 34 | 0 | 28 | 2 | 14 | 2 | 28 | 2 | 34 | 0 | 30 | 1 |  |  |  |  |  |  |
|  | result (n) | + | 6 | 56 | 13 | 49 | 11 | 41 | 7 | 49 | 11 | 41 | 11 | 51 | 9 | 47 |  |  |  |  |  |  |
|  | Cohen's Kappa |  | 0.848 |  | 0.728 |  | 0.679 |  | 0.675 |  | 0.679 |  | 0.767 |  | 0.763 |  |  |  |  |  |  |  |
|  | 95% Lower |  | 0.758 |  | 0.625 |  | 0.565 |  | 0.553 |  | 0.565 |  | 0.667 |  | 0.658 |  |  |  |  |  |  |  |
|  | CI Upper |  | 0.910 |  | 0.811 |  | 0.775 |  | 0.778 |  | 0.775 |  | 0.844 |  | 0.845 |  |  |  |  |  |  |  |
| Getein | Test | - | 33 | 6 | 42 | 2 | 37 | 4 | 16 | 4 | 37 | 4 | 42 | 2 |  |  |  |  |  |  |  |  |
|  | result (n) | + | 3 | 45 | 8 | 45 | 6 | 43 | 6 | 46 | 6 | 43 | 6 | 47 |  |  |  |  |  |  |  |  |
|  | Cohen's Kappa |  | 0.789 |  | 0.794 |  | 0.777 |  | 0.664 |  | 0.777 |  | 0.835 |  |  |  |  |  |  |  |  |  |
|  | 95% Lower |  | 0.686 |  | 0.698 |  | 0.675 |  | 0.542 |  | 0.675 |  | 0.743 |  |  |  |  |  |  |  |  |  |
|  | CI Upper |  | 0.867 |  | 0.867 |  | 0.855 |  | 0.769 |  | 0.855 |  | 0.900 |  |  |  |  |  |  |  |  |  |
| Innovita | Test | - | 38 | 7 | 55 | 1 | 42 | 3 | 25 | 7 | 42 | 3 |  |  |  |  |  |  |  |  |  |  |
|  | result (n) | + | 1 | 50 | 5 | 59 | 2 | 44 | 5 | 67 | 2 | 44 |  |  |  |  |  |  |  |  |  |  |
|  | Cohen's Kappa |  | 0.831 |  | 0.900 |  | 0.890 |  | 0.724 |  | 0.890 |  |  |  |  |  |  |  |  |  |  |  |
|  | 95% Lower |  | 0.738 |  | 0.828 |  | 0.803 |  | 0.627 |  | 0.803 |  |  |  |  |  |  |  |  |  |  |  |
|  | CI Upper |  | 0.897 |  | 0.945 |  | 0.943 |  | 0.805 |  | 0.943 |  |  |  |  |  |  |  |  |  |  |  |
| Leccurate | Test | - | 37 | 7 | 42 | 1 | 37 | 2 | 15 | 5 |  |  |  |  |  |  |  |  |  |  |  |  |
|  | result (n) | + | 3 | 50 | 5 | 48 | 2 | 41 | 6 | 46 |  |  |  |  |  |  |  |  |  |  |  |  |
|  | Cohen's Kappa |  | 0.790 |  | 0.875 |  | 0.902 |  | 0.625 |  |  |  |  |  |  |  |  |  |  |  |  |  |
|  | 95% Lower |  | 0.694 |  | 0.788 |  | 0.811 |  | 0.503 |  |  |  |  |  |  |  |  |  |  |  |  |  |
|  | CI Upper |  | 0.864 |  | 0.931 |  | 0.954 |  | 0.734 |  |  |  |  |  |  |  |  |  |  |  |  |  |
| Liming | Test | - | 15 | 1 | 26 | 8 | 17 | 4 |  |  |  |  |  |  |  |  |  |  |  |  |  |  |
|  | result (n) | + | 6 | 50 | 4 | 56 | 5 | 41 |  |  |  |  |  |  |  |  |  |  |  |  |  |  |
|  | Cohen's Kappa |  | 0.747 |  | 0.716 |  | 0.692 |  |  |  |  |  |  |  |  |  |  |  |  |  |  |  |
|  | 95% Lower |  | 0.628 |  | 0.612 |  | 0.566 |  |  |  |  |  |  |  |  |  |  |  |  |  |  |  |
|  | CI Upper |  | 0.839 |  | 0.802 |  | 0.796 |  |  |  |  |  |  |  |  |  |  |  |  |  |  |  |
| Medomics | Test | - | 32 | 7 | 44 | 0 |  |  |  |  |  |  |  |  |  |  |  |  |  |  |  |  |
|  | result (n) | + | 2 | 41 | 4 | 43 |  |  |  |  |  |  |  |  |  |  |  |  |  |  |  |  |
|  | Cohen's Kappa |  | 0.779 |  | 0.912 |  |  |  |  |  |  |  |  |  |  |  |  |  |  |  |  |  |
|  | 95% Lower |  | 0.671 |  | 0.829 |  |  |  |  |  |  |  |  |  |  |  |  |  |  |  |  |  |
|  | CI Upper |  | 0.860 |  | 0.959 |  |  |  |  |  |  |  |  |  |  |  |  |  |  |  |  |  |
| Render | Test | - | 39 | 8 |  |  |  |  |  |  |  |  |  |  |  |  |  |  |  |  |  |  |
|  | result (n) | + | 0 | 49 |  |  |  |  |  |  |  |  |  |  |  |  |  |  |  |  |  |  |
|  | Cohen's Kappa |  | 0.833 |  |  |  |  |  |  |  |  |  |  |  |  |  |  |  |  |  |  |  |
|  | 95% Lower |  | 0.740 |  |  |  |  |  |  |  |  |  |  |  |  |  |  |  |  |  |  |  |
|  | CI Upper |  | 0.898 |  |  |  |  |  |  |  |  |  |  |  |  |  |  |  |  |  |  |  |

Four out of 10 tests, and 5 out of 9 tests have positive predictive values (PPV) of 100% for the detection of IgG and IgA/IgM, respectively (Supplementary Table S5). Considering the combined tests, 2 of them predict positive cases with 100% accuracy (Liming and Render); Cellex has the lowest PPV (90.2%). Euroimmun test (IgG + IgA) shows the best negative predictive value (NPV), correctly classifying a negative case in 68.5% of the cases tested. Focusing on each test independently, Abbott performs the best in identifying the negative cases based on IgG detection (57.8%) and SD based on IgM detection (61.0%); however, Euroimmun IgA is the test with the highest NPV (66.1%; Supplementary Table S5).

Analysis of the ROC curves of the semiquantitative tests reveals that Euroimmun is the test that best distinguishes SARS-CoV-2 non-infected controls from the SARS-CoV-2 confirmed cases, both for IgA and IgG [AUC (IgG) = 0.911; AUC (IgA) = 0.935; Figure 2].

**Figure 2:**
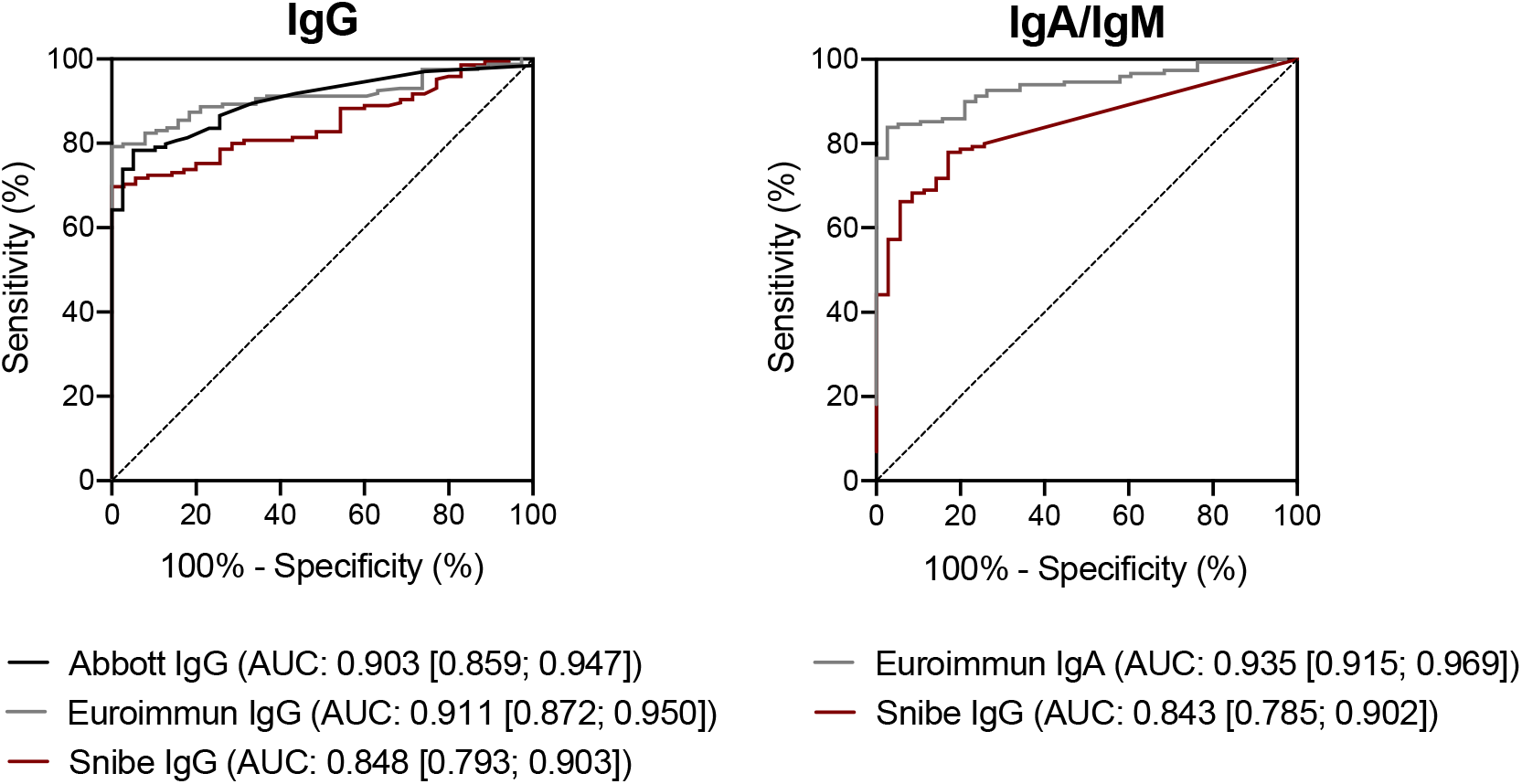
Receiver operator characteristic (ROC) curves of the assayed semiquantitative tests. AUC: area under the curve.

### Comparison of immunoglobulins levels in COVID-19 patients with distinct clinical presentation

Overall, all the Ig assayed start being detected at early days since symptoms onset (<10 days). IgM and IgG reach the maximum relative amount at 16-20 days or at >21days. A peak of IgA detection is observed at 16-21 days since symptoms onset (Supplementary Figure S2).

To further investigate the association of Ig levels with clinical presentation, COVID-19 inpatients were classified as severe or non-severe, based on the need of supplementary oxygen or mechanic ventilatory support, as specified in the Methods section. The relative amount of Ig detected on each of the semiquantitative tests was compared between the two subgroups (Figure 3). IgG relative amounts tend to be lower in the severe group at <10 days when compared with the non-severe group, reaching statistical significance for Abbot and Euroimmun [Abbott: U = 7.5, *p* = 0.0006; Euroimmun IgG: U = 23.5, *p* = 0.0045]. Accordingly, Euroimmun IgA and Snibe IgM also present lower relative amounts in the severe group at <10 days post-symptoms onset when compared with the non-severe group [Euroimmun IgA: U = 29.5, *p* = 0.0133; Snibe IgM: U = 33.5, *p* = 0,0446]. For Snibe, both IgM and IgG levels at >21 days are higher in the severe group [Snibe IgM: U = 145.5, *p* = 0.0106; Snibe IgG: U = 110, p = 0.0064]. No differences are observed in the other tests or time ranges analysed (Figure 3).

**Figure 3:**
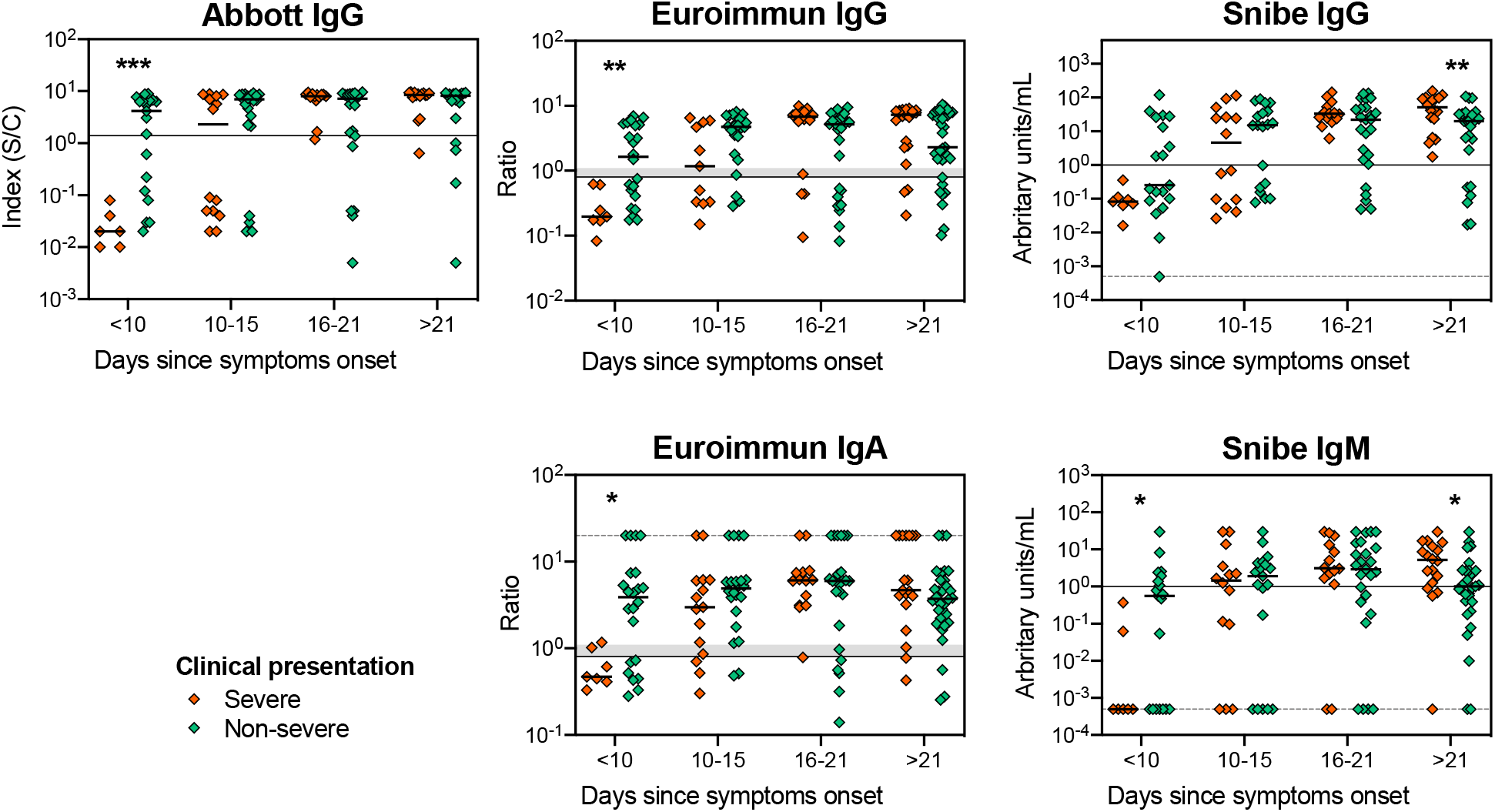
Comparison of the Ig levels in patients with severe and non-severe clinical presentation using the semiquantitative tests Abbott, Euroimmun and Snibe. Each dot represents one sample, and the solid thick lines correspond to group’s median. Solid thin lines represent each test’s cut-off value. In the case of Euroimmun test, the shadowed grey banner refers to borderline values according to the manufacturer (between 0.8 and 1.1). Since the y-axis has a log scale that does not allow representation of zero, in those situations (Snibe IgG and IgM) arbitrary values were attributed, and a dashed grey line was represented in this value. For values above the detection limit (Euroimmun IgA), a random value of 20 was attributed and represented as a dashed grey line. Groups median were compared using a MannWhitney U-tests; significant differences were represented as: * for *p* < 0.05; ** for *p* < 0.01; ***, *p* < 0.001.

## DISCUSSION

Serological testing to SARS-CoV-2 infection is a rapid, inexpensive, and easy diagnostic approach complementary to RT-qPCR, but also an essential tool to access the presence and profile of Ig anti-SARS-CoV-2, both individually and at the population level. The present study compares the performance of serological tests towards SARS-CoV-2 using well characterized COVID-19 inpatients at various moments of the disease course, and with diverse clinical presentation severity. This study is relevant since most commercially available tests emerged in the market soon after the COVID-19 outbreak and were based on the analysis of a limited number of patients. In the recent months, many studies exploring the production of Ig against SARS-CoV-2 have been reported, but few compare several tests between them using the same set of plasma samples. Comparison of tests performance, as presented here, is of relevance since it depends on inherent population genetic variations, on time during course of the disease and on disease severity.

Irrespective of the test methodology (ELISA, CLIA, LFIA), we observe here, as reported previously, that test sensitivity is dependent on time since symptoms onset. In addition, the combined detection of IgG and IgA/IgM against SARS-CoV-2 leads, in most cases, to a better performance than measurements of a single Ig class, regardless of the time range evaluated. These observations were previously reported by others, and are in agreement with the establishment of the immune response, where IgA, IgM and IgG have different dynamics throughout disease progression (5,12-18, 25). This dynamic nature of tests sensitivity needs to be considered when interpreting performance. Inconsistent reporting by manufactures, or lack of details about the timepoints used to establish tests’ performance, caused confusion about the expected sensitivity. This has contributed to the recent decision by the US Food and Drug Administration to remove some tests from the Emergency Use Authorization.

Previous reports concluded that N-protein/peptide-based tests present better sensitivities than the ones based on S-protein/peptide. This difference may result from an earlier immune response towards the N-protein/peptide in comparison to the one towards the S-protein/peptide, or related to the higher specificity of Ig towards the N-protein/peptide (26-28). It is not possible to confront these previous results with ours, as 4 of the assayed tests displayed no information regarding the target antigen, 4 target both proteins, 2 target only the N-protein/peptide alone; and 1 target only the S-protein.

Regarding seroconversion rate, it is striking to notice that Ig anti-SARS-CoV-2 are detectable at very early days since symptoms onset (at 3 to 5 days either for IgM, IgA or IgG; *e.g*. Patients 012, 033 and 070; Supplementary Table S2), though for others, detection using the same tests occurs only much later (>21 days; *e.g*. Patients 004, 037 and 064; Supplementary Table S2). Several factors might account for these observations. Antibody production kinetics during SARS-CoV-2 infection is not yet fully elucidated, nor are the factors responsible for differences in patients’ response. As others suggested previously (17,18), we also observe that patients with non-severe clinical presentation show higher relative amounts of IgG, IgA and IgM at the earlier period since symptoms onset. Identifying the variable responsible for distinct profiles of seroconversion is necessary to be able to fully understand false-negative results.

It is important to recall that the time since symptoms onset is self-reported, which might introduce variability taking into consideration the diversity of clinical manifestation of COVID-19 and of symptoms perception by each individual. However, we consider that it introduces less variability than time since disease diagnosis, which is performed for some patients before symptoms onset and for others several days after.

Due to the high specificity of most of the assayed tests, their PPV are relatively high (over 90%). Individuals with positive serodiagnosis tests have a high likelihood of being infected by SARS-CoV-2 (past or present infection). However, a negative serologic test in these settings should not be interpreted as not being infected with SARS-CoV-2 as the NPV are relatively low (38% to 68%).

Our results highlight that Ig levels, on a hospitalized COVID-19 population, depend on disease severity. In fact, our data suggests a delay in Ig detection in patients with worse disease presentation; they present lower relative amounts of Ig at the initial phase of the disease and higher at later stages, in comparison to patients with non-severe clinical presentation. This information is of outmost relevance in clinical settings and needs to be further explored in a larger cohort of patients evaluated longitudinally, since the very beginning of symptoms manifestation perception, and throughout longer time-periods.

Our results are based on measurements performed in a specific population of hospitalized COVID-19 patients and should not be extrapolated to the general population. Still, it provides the basis for an informed selection of a serological test to be assayed and applied in a greater population setting to evaluate the potential value of each test as a complement for COVID-19 diagnosis and to understand the dynamics of Ig production upon infection with SARS-CoV-2.

## Data Availability

All data generated or analysed during this study are included in this published article and its supplementary information file.

## ACKNOWLEDGMENTS

This work was funded by National funds, through the Foundation for Science and Technology (FCT): R4COVID (596694995), POCI-01-0145-FEDER-016428, UIDB/50026/2020 and UIDP/50026/2020; and by the projects NORTE-01-0145-FEDER-000013 and NORTE-01-0145-FEDER-000023, supported by Norte Portugal Regional Operational Programme (NORTE 2020), under the PORTUGAL 2020 Partnership Agreement, through the European Regional Development Fund (ERDF). CN, SR and NV are junior researchers under the scope of the FCT Transitional Rule DL57/2016. JC-G is supported by a FCT PhD grant, in the context the Doctoral Program in Aging and Chronic Diseases (PhDOC; PD/BD/137433/2018); CSS is supported by a FCT PhD grant, in the context of the Doctoral Program in Applied Health Sciences (PD/BDE/142976/2018). The authors gratefully acknowledge Drs. AC Braga and R Menezes (University of Minho) for discussions of the statistical properties of the data, Dr. Qi Huan (Nanjing Tembusu New Material Research Institute) for assistance in obtaining the Medomics kits, Dr. Lei Wu (INL) for assistance in obtaining the Getein kits and Hao Tu (Overseas Business, Getein) for providing the Getein kits for this project free of charge. We are thankful to all study participants.

## AUTHOR’S CONTRIBUTION

CS-M, CN, SR, CC, JAP, PGC and MC-N conceptualized the study; CS-M, CN, SR and MC-N designed the experiments; JC, AR, MF, HS, OP, AC, CC and PGC recruited the patients and collected the clinical data; CS-M, CN, SR, JC-G, CSS, NV, PB-S and PA-P performed the experiments; CS-M, CN and SR prepared the figures and performed the statistical analysis; JAP, PGC and MC-N supervised the study; and all authors discussed the results and contributed to the final manuscript.

